# Schizophrenia polygenic risk score and cannabis use modify psychosis expression in first episode psychosis patients and population controls

**DOI:** 10.1101/19013284

**Authors:** Diego Quattrone, Ulrich Reininghaus, Alex L. Richards, Giada Tripoli, Laura Ferraro, Paolo Marino, Victoria Rodriguez, EU-GEI group, Charlotte Gayer-Anderson, Hannah E. Jongsma, Peter B. Jones, Caterina La Cascia, Daniele La Barbera, Ilaria Tarricone, Elena Bonora, Sarah Tosato, Antonio Lasalvia, Andrei Szöke, Celso Arango, Miquel Bernardo, Julio Bobes, Cristina Marta Del Ben, Paulo Rossi Menezes, Pierre-Michel Llorca, Jose Luis Santos, Julio Sanjuán, Andrea Tortelli, Eva Velthorst, Lieuwe de Haan, Bart P.F. Rutten, Michael T. Lynskey, Tom P. Freeman, James B. Kirkbride, Pak C. Sham, Michael C. O’Donovan, Alastair Cardno, Evangelos Vassos, Jim van Os, Craig Morgan, Robin M. Murray, Cathryn M. Lewis, Marta Di Forti

## Abstract

**Background:** Diagnostic categories within the psychosis spectrum are widely used in clinical practice, however psychosis may occur on a continuum. Therefore, we explored whether the continuous distribution of psychotic symptoms across categories is a function of genetic as well as environmental risk factors, such as polygenic risk scores (PRSs) and cannabis use.

**Methods:** As part of the EU-GEI study, we genotyped first episode psychosis patients (FEP) and population controls, for whom transdiagnostic dimensions of psychotic symptoms or experiences were generated using item response bi-factor modelling. Linear regression was used, separately in patients and controls, to test the associations between these dimensions and schizophrenia (SZ) PRSs, as well as the combined effect of SZ-PRS and cannabis use on the positive symptom/experience dimensions.

**Results:** SZ-PRS was associated with negative (B=0.18; 95%CI 0.03 to 0.34) and positive (B=0.19; 95%CI 0.03 to 0.36) symptom dimensions in 617 FEP, and with all the psychotic experience dimensions in 979 controls. The putative effect of SZ-PRS on either symptom or experience dimensions was of a small magnitude. Cannabis use was additionally associated with the positive dimensions both in FEP (B=0.31; 95%CI 0.11 to 0.52) and in controls (B=0.26; 95%CI 0.06 to 0.46), independently from SZ-PRS.

**Conclusions:** We report two validators to the latent dimensional structure of psychosis. SZ risk variants and cannabis use independently map onto specific dimensions, contributing to variation across the psychosis continuum. Findings support the hypothesis that psychotic experiences have similar biological substrates as clinical disorders.

## Introduction

Psychotic disorders are syndromes caused by multiple genetic and socioenvironmental factors (1). However, the current classification system is based on a ‘natural history approach’ rather than on a ‘natural classification’ (2). Specifically, diagnostic categories of non-affective (e.g., schizophrenia, schizoaffective disorders) and affective (e.g., bipolar disorder, psychotic depression) psychosis were developed from observed similarities and dissimilarities of signs and symptoms over time, without considering biological or socio-environmental factors (3). Hence, the question of whether current diagnostic categories are the most valid phenotypes for research is still debated, due to the following methodological limitations.

First, psychotic disorders are commonly studied as binary phenotypes (e.g., diagnosis yes/no), although psychotic symptoms follow a continuous distribution (4). Furthermore, some authorities claim that the introduction of operationalised classification systems in the 1970s led to the ‘death of phenomenology’ driving biological psychiatry to focus on the presence/absence of a diagnosis whilst overlooking the complex expression of psychotic phenomena (5). Moreover, Kraepelin’s paradigm (i.e., the neat distinction between non-affective and affective psychosis) has been challenged (6), though not yet replaced (3). As a consequence, the high comorbidity indices among psychotic disorders (7), as well as their high genetic correlation (8), may be an artefact of our own diagnostic conceptualization.

To address these limitations, the use an approach based on symptom dimensions has been proposed (9). Consistent with this methodology, we reported that transdiagnostic psychopathology at first episode psychosis (FEP) can be represented by a general psychosis factor (G), and five specific dimensions of positive (POS), negative (NEG), disorganization (DIS), manic (MAN), and depressive (DEP) symptoms (10). Similarly, a model composed of general and specific experience dimensions has been proposed to measure subclinical psychosis in the general population (11, 12). These conceptualizations statistically reflect a ‘bi-factor model’, where the general and specific dimensions account, respectively, for the unidimensional and multidimensional nature of the latent psychosis construct (13, 14). We have previously advocated that such structures should be validated by the degree to which biological and environmental factors cohere with general and specific symptom dimensions (10). Indeed, according to the coherence theory of truth, psychiatric constructs can be approximated as true if they are well connected and integrated into our accumulated scientific evidence (15).

Thus, we recently found evidence that cannabis-associated psychopathology at psychosis onset is characterised by high POS scores and low NEG scores (16). In relation to biological factors, symptom dimensions have been investigated in family, twin and adoption studies (17-22), overall showing that NEG or DIS symptoms had higher familial aggregation than other symptom dimensions.

In recent years the availability of summary statistics from large genome-wide association studies (GWAS) across psychiatric phenotypes has allowed researchers to test in independent samples how the genetic liability to a disorder predicts any other traits (23). Genetic liability is commonly summarised into a polygenic risk score (PRS) (24), however, only a few studies to date have investigated the relation between SZ-PRS and psychotic symptom dimensions (25). In patients, an association between SZ-PRS and NEG (or DIS) symptoms was found in several SZ studies (26-28) and in Psychiatric Genomics Consortium (PGC) large mega-analyses (29, 30). However, other studies have not found the same pattern of associations (31, 32), and only one study reported that SZ-PRS correlated with POS symptoms (28). Interestingly, in the general population an association was observed between SZ-PRS and either NEG (12, 33) or POS psychotic experiences (34-36); however, negative findings have also been reported (37).

The inconsistency across studies could be explained by differences in study design, methods, and GWAS power. Of note, only one small study examined a FEP sample (38), in which confounding effects of antipsychotic drugs on symptoms are minimised and a common comparable time point in the course of illness is used. In addition, most studies have not performed factor analysis of observed symptoms to measure and validate latent constructs. Finally, no studies have applied summary statistics from recent PGC GWAS investigating similarities and dissimilarities between SZ and BP (29).

We have previously reported findings from bi-factor models of 1) psychotic symptoms in a multinational FEP sample (10) and 2) psychotic experiences in controls representative of the population at risk in each catchment area (11). In the current study, we aimed to investigate the association between these phenotypes and genetic loading for SZ and BP, as summarised by i) SZ-PRS, ii) BP-PRS, iii) combined SZ *&* BP-PRS. We further explored whether the previously reported association of cannabis use with the POS dimensions (16) holds when taking into account SZ-PRS.

Based on an *a priori* synopsis, we hypothesized that SZ-PRS would be positively associated with the NEG dimension in FEP patients, and with the POS dimensions in both FEP patients and the general population. Furthermore, we hypothesized a cumulative effect of cannabis use on POS dimensions independent of SZ-PRS. Finally, we expected in FEP, an association between BP-PRS and the MAN symptom dimension, and between the combined SZ & BP-PRS and the G factor.

## Methods and Materials

### Sample design and procedures

FEP patients and population controls were recruited as part of the EUropean network of national schizophrenia networks studying Gene-Environment Interactions (EU-GEI). FEP patients were identified between 2010 and 2015 across six countries to examine incidence rates of psychotic disorders and patterns of symptomatology (10, 39). For examining biological and environmental risk factors, DNA samples were collected, and an extensive face-to-face assessment was conducted on 1,130 FEP and 1,497 controls, broadly representative of the population living in each catchment area by age, sex and ethnic group. More information on recruitment strategies is available in earlier EU-GEI incidence and case-control papers (39, 40).

### Measures

Data on age, sex, and ethnicity were collected using a modified version of the Medical Research Council Sociodemographic Schedule (41).

The OPerational CRITeria (OPCRIT) system (42) was used by centrally trained investigators, whose reliability was assessed before and throughout the study (k=0.7), to assess psychopathology experienced in the first four weeks after FEP and define research-based diagnoses. Moreover, psychopathology assessment included the use of the Schedule for Deficit Syndrome (SDS) (43) to evaluate NEG symptoms, which are not extensively covered by the OPCRIT.

The Community Assessment of Psychic Experiences (CAPE) (44) was administered to population controls to report their positive, negative, and depressive psychotic experiences. A modified version of the Cannabis Experience Questionnaire (CEQ_EU-GEI_) (45) was used to collect extensive information on patterns of cannabis use.

### Dimensions of psychotic symptoms and experiences

Data from OPCRIT and CAPE were analysed using item response modelling in M*plus*, version 7.4, to estimate two bifactor models of psychopathology, based on the associations among observer ratings of psychotic symptoms in patients or self-rating of psychotic experiences in controls (Supplementary Figures S1 and S2). This methodology is described in full in earlier EU-GEI papers on transdiagnostic dimensions (10, 11). Briefly, OPCRIT or CAPE items were dichotomized as 0 ‘absent’ or 1 ‘present’, to estimate two bi-factor models, for patients and controls respectively. Bi-factor solutions were compared with competitive solutions (i.e., unidimensional, multidimensional, hierarchical models of psychosis) using Log-Likelihood (LL), Akaike Information Criterion (AIC), Bayesian Information Criterion (BIC), and Sample-size Adjusted BIC (SABIC) as model fit statistics. McDonald’s omega (ω) (46), omega hierarchical (ω_H_) (46), and index *H* (47), were computed as reliability and strength indices.

Data from SDS were analysed in M*plus*, version 7.4, following the same procedure as described above. We did not estimate a bi-factor model for SDS due to lack of rationale for a G factor of negative symptoms. Instead, based on the structure of the NEG construct (48) and previous factor analysis studies on SDS (49), we estimated a multidimensional model of NEG symptoms composed of the two specific dimensions of 1) ‘avolition’ and 2) (lack of) ‘emotional expressivity’. We considered ‘emotional expressivity’ as the most genuine phenotypic expression of primary negative symptoms for subsequent analysis, as the behavioural manifestation of ‘avolition’ may partly overlap with depressive symptoms in a FEP sample. SDS was not administered in one of the study sites, Verona, which was therefore not included in the analysis of NEG symptoms.

### Genotype procedure

The EU-GEI case-control sample was genotyped at the MRC Centre for Neuropsychiatric Genetics and Genomics in Cardiff (UK) using a custom Illumina HumanCoreExome-24 BeadChip genotyping array covering 570,038 genetic variants. Imputation was performed in the Michigan Imputation Server, using the Haplotype Reference Consortium reference panel, with Eagle software for estimating haplotype phase, and Minimac3 for genotype imputation (50-52). The imputed best-guess genotype was used for the present analysis.

### Population stratification and polygenic risk score calculation

We performed a two-step procedure to account for the multi-ethnic nature of the sample (reported in full in the supplementary material), by excluding populations in our sample of very different ancestry from external GWAS data. Briefly, as a first step, we identified in our sample ancestry clusters of individuals through iterative pruning of principal component analysis (ipPCA) of single nucleotide polymorphisms (SNPs), and we tested for each cluster whether PRS discriminated cases from controls. As a second step, we merged these clusters (based on whether PRS had discriminative value), removed long-range genome regions with complex linkage disequilibrium (LD) patterns, recalculated main Principal Components (PCs), and finally constructed main PRSs using PRSice (53). Specifically, individuals’ risk variants were weighted by the log(odds ratio), where the odds ratio was extracted from the latest summary statistics of SZ and BP PGC mega-analyses (29, 54, 55), which did not include any EU-GEI sample. Logistic regression was then applied to predict case status from SZ- and BP-PRS, after covarying for 10 PCs, sex, age, and primary diagnosis. Nagelkerke’s *R*^*2*^ was used as a measure of the difference in variance between the full-model versus a model with the covariates alone, at the SNPs p-value threshold (*P*_*T*_)=0.05 (selected *a priori* as it maximised the explained variance in case status in the PGC studies (54, 55)).

### Relationship between symptom dimensions, polygenic risk scores, and cannabis use

We tested for associations between PRSs and the scores on transdiagnostic dimensions of psychotic symptoms/experiences, separately in FEP and controls, using linear regression.

Specifically, in FEP we tested for association between all symptom dimensions and the three PRSs. In controls, we tested for association between all psychotic experience dimensions and SZ-PRS; we did not test BP-related PRSs since (hypo)manic experiences were not rated in our controls.

Moreover, we used predicted values of SZ-PRS after regression of case/control status, to illustrate the continuous distribution of SZ-PRS in our sample according to quartiles of positive psychotic experience and symptoms.

To examine the combined associations of cannabis use and SZ-PRS with POS dimensions, we selected the two variables on pattern of cannabis use previously associated with POS (11), i.e., ‘lifetime daily use’ in patients and ‘current use’ in controls. We first checked for correlation with SZ-PRS, and subsequently we added the two cannabis terms to the models.

Given the high number of outcomes (six dimensions in patients, four in controls) and predictors (PRSs and cannabis use), and the number of hypotheses (four in patients, one in controls), we controlled the false discovery rate using the Benjamini and Hochberg procedure (56), tolerating a 10% false discovery rate (*q*=0.10).

Furthermore, as a sensitivity measure, in PRSice we tested whether the main effect of PRSs on dimensions held at other P_T_ and ran a permutation analysis to further control the familywise error rate, by repeating the PRSice procedure shuffling the phenotype 5,000 times to obtain an empirical distribution of the p-value at the best *P*_*T*_. Finally, we used AVENGEME (Additive Variance Explained and Number of Genetic Effects Method of Estimation) to further evaluate the consistency of the effect directions across different *P*_*T*_ and compute the genetic covariance (σ12) between our symptom dimensions and the PGC GWAS data (57).

## Results

### Main PCs and PRS computation

Population stratification findings are presented in the supplement material. Based on the case-control discriminative value of SZ- and BP-PRS in each population cluster, we merged 1,596 individuals (617 FEP and 979 population controls) for SZ-PRS analyses, and 505 FEP for BP-PRS analysis only. The ability of SZ and BP PRSs to distinguish cases from controls in the main sample is presented in Figure 1, showing that at P_T_=0.05, SZ-PRS accounted for a Nagelkerke’s R^2^ of 0.09 (p=6.9×10^−26^); and BP-PRS for a Nagelkerke’s R^2^ of 0.02 (p=5×10^−6^).

**Figure 1.**
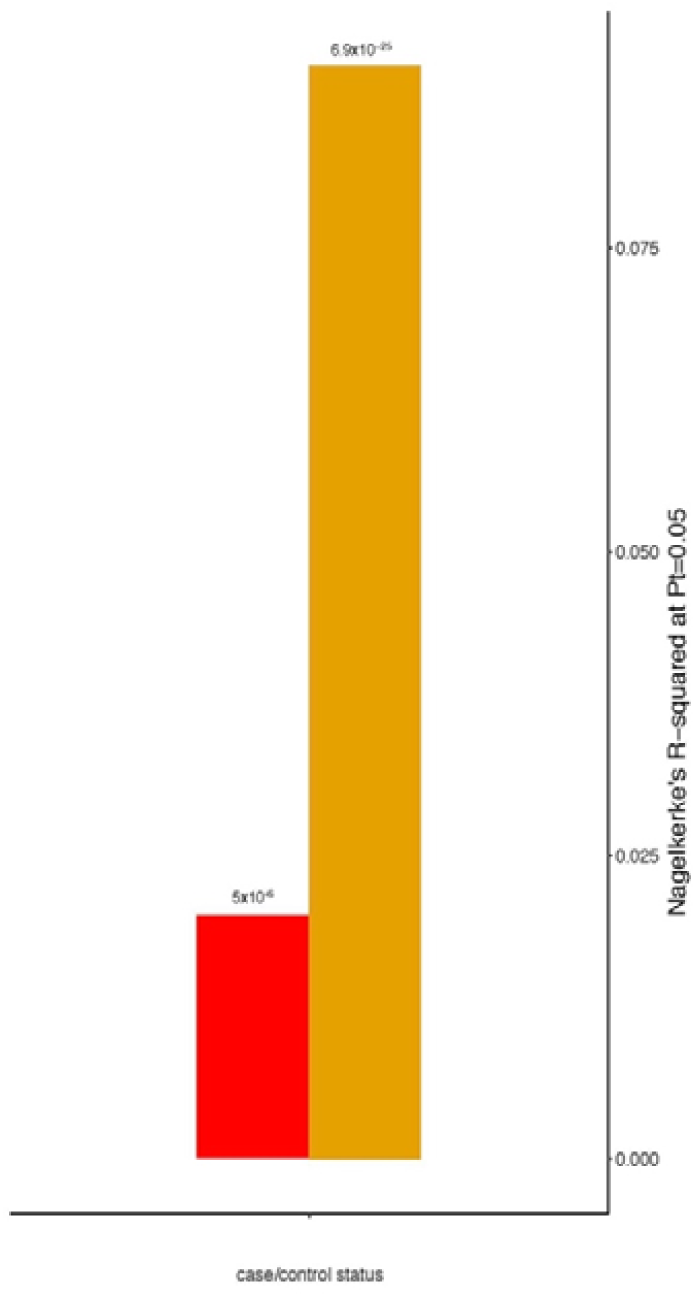
SZ-PRS and BP-PRS by FEP-control status. The bar plot shows the variance in case-control status (y-axis) explained by SZ-PRS (yellow) and BP-PRS (red) respectively. P-value is presented on top of the bars.

### Psychotic symptom dimensions by PRS in patients

Findings on symptom dimensions in cases by SZ-, BP-, and SZ *&* BP-PRSs at P_T_=0.05 are shown in Table 1 and Figure 1. As expected in PRS cross-trait predictions (23), the magnitude of the SNPs effect was small for all the associations detected. Specifically, SZ-PRS was associated with a high score for both the positive (B=0.19, 95% CI 0.03 to 0.35; Nagelkerke’s R^2^ =0.009, p=0.019) and negative (B=0.18, 95% CI 0.03 to 0.33; Nagelkerke’s R^2^ =0.01, p=0.021) symptom dimensions. Moreover, we found no nominal association between BP-PRS and the MAN symptom dimension (B=0.09, 95% CI −0.01 to 0.19; Nagelkerke’s R^2^ =0.008, p=0.055); and between SZ *&* BP-PRS and the G factor (B=0.06, 95%CI −0.05 to 0.16; p=0.158).

**Table 1.** Symptom dimension scores by PRSs in cases.

|  | General<sup>a</sup><br>B (95% CI) | Positive<sup>a</sup><br>B (95% CI) | Negative<sup>b</sup><br>B (95% CI) | Disorganization<sup>a</sup><br>B (95% CI) | Mania<sup>a</sup><br>B (95% CI) | Depression<sup>a</sup><br>B (95% CI) |
| --- | --- | --- | --- | --- | --- | --- |
| **SZ PRS model** | 0.04<br>(-0.09 to 0.18)<br>p=0.528 | 0.19<br>(0.03 to 0.35)<br>**p=0.021\*<sup>†</sup>** | 0.16<br>(0.1 to 0.3)<br>**p=0.019\*<sup>†</sup>** | -0.01<br>(-0.16 to 0.14)<br>p=0.928 | 0.06<br>(-0.07 to 0.2)<br>p=0.378 | -0.06<br>(-0.2 to 0.07)<br>p=0.350 |
| **BP PRS model** | 0.06<br>(-0.03 to 0.15)<br>p=0.175 | 0.05<br>(-0.06 to 0.17)<br>p=0.341 | -0.005<br>(-0.09 to 0.08)<br>p=0.915 | 0.01<br>(-0.1 to 0.1)<br>p=0.976 | 0.09<br>(-0.01 to 0.19)<br>p=0.055 | -0.01<br>(-0.1 to 0.08)<br>p=0.938 |

Explanatory note. B, Unstandardized regression coefficient; CI, confidence interval. Covariates in multiple models were sex, age, ten ancestry PCs, and categorical diagnosis.
a. Symptom dimension scores from OPCRIT factor analysis.
b. Symptom dimension scores from SDS factor analysis.
Associations nominally significant after permutation analysis are showed in bold
\*P-values nominally significant after Benjamini-Hochberg procedure at FDR threshold = 0.1
(<sup>†</sup>Benjamini-Hochberg P-value: 0.056)

Sensitivity analysis showed that the pattern of associations between SZ-PRS with either POS or NEG symptom dimensions was consistently observed across all P_T_ and remained relevant even after permutation analysis (Figure S6 – supplement, showing empirical p-values of 0.007 for POS; and of 0.055 for NEG). Furthermore, a positive genetic covariance was observed between both NEG and PGC SZ GWAS [σ12=0.56 (95%CI 0.39 to 0.76)] and POS and PGC SZ GWAS [σ12=0.51 (95%CI 0.35 to 0.69)].

Finally, the violin plots presented in Figure 2 illustrate the kernel distribution of predicted value of SZ-PRS across individual quartiles of positive psychotic symptoms.

**Figure 2.**
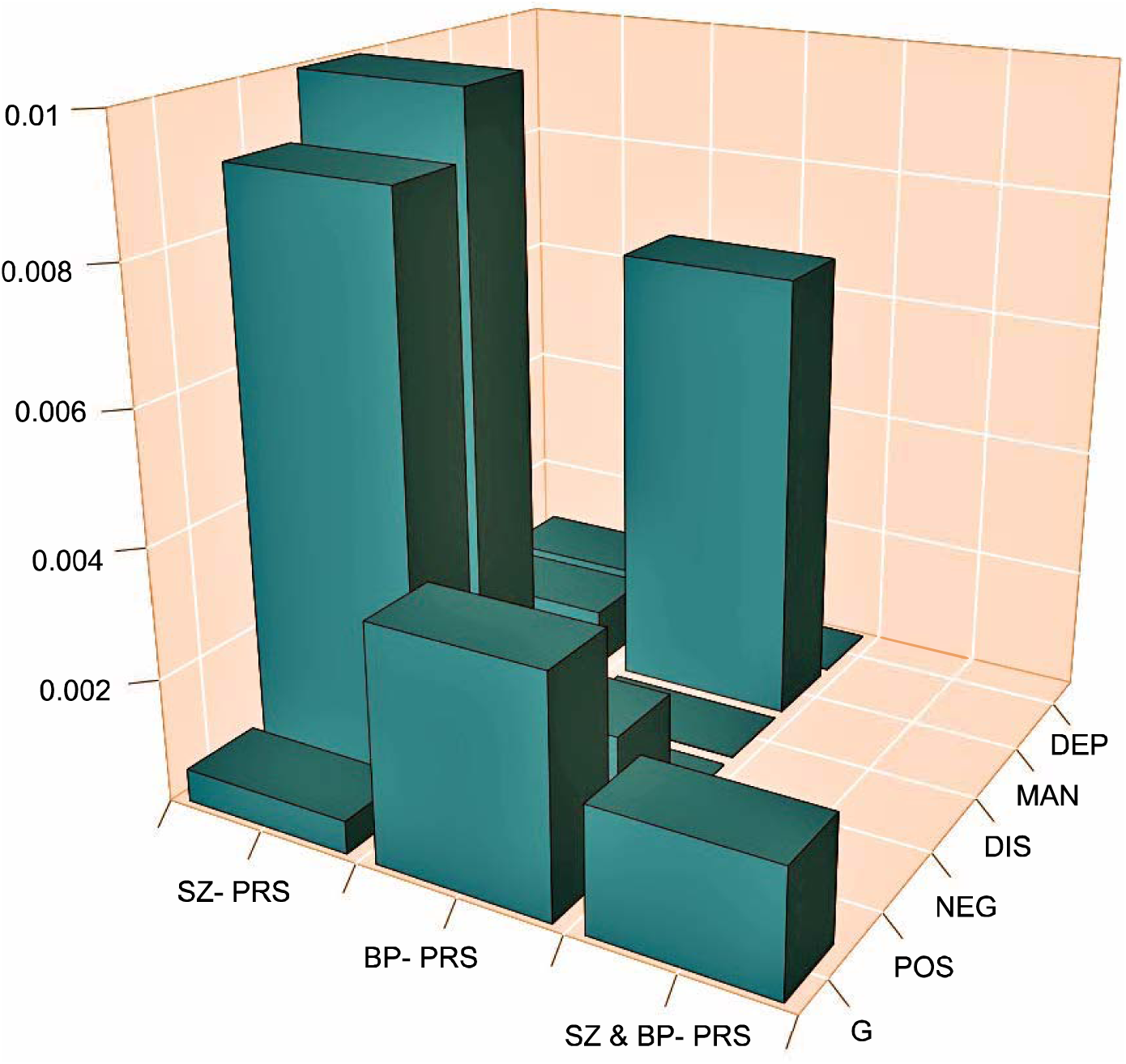
SZ- PRS, BP- PRS, and SZ & BP- PRS by symptom dimensions in FEP. The bar plot shows the variance (y-axis, Nagelkerke’s *R*^*2*^) explained by the different PRSs (x-axis) for each symptom dimension (z-axis).

### Psychotic experience dimensions by SZ-PRS in controls

A positive association between SZ-PRS with a higher score at all psychotic experience dimensions was found (Table 2). Sensitivity analysis showed that the association between SZ-PRS with POS psychotic experiences was consistent across different *P*_*T*_ and remained relevant after permutation analysis (Figure S7 – supplement, showing an empirical P-value = 0.003). The kernel distribution of predicted value of SZ-PRS according to individual quartiles of psychotic experiences in controls is reported in Figure 3.

**Table 2.**
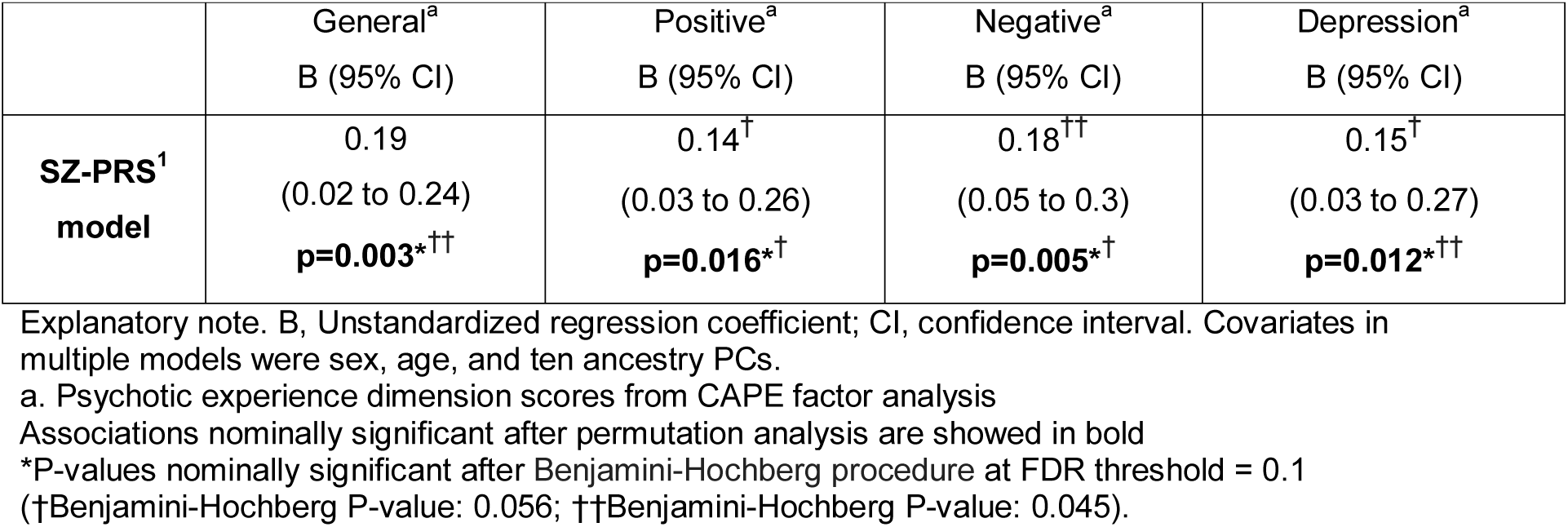
Psychotic experience dimension scores by SZ-PRS in controls.

**Figure 3.**
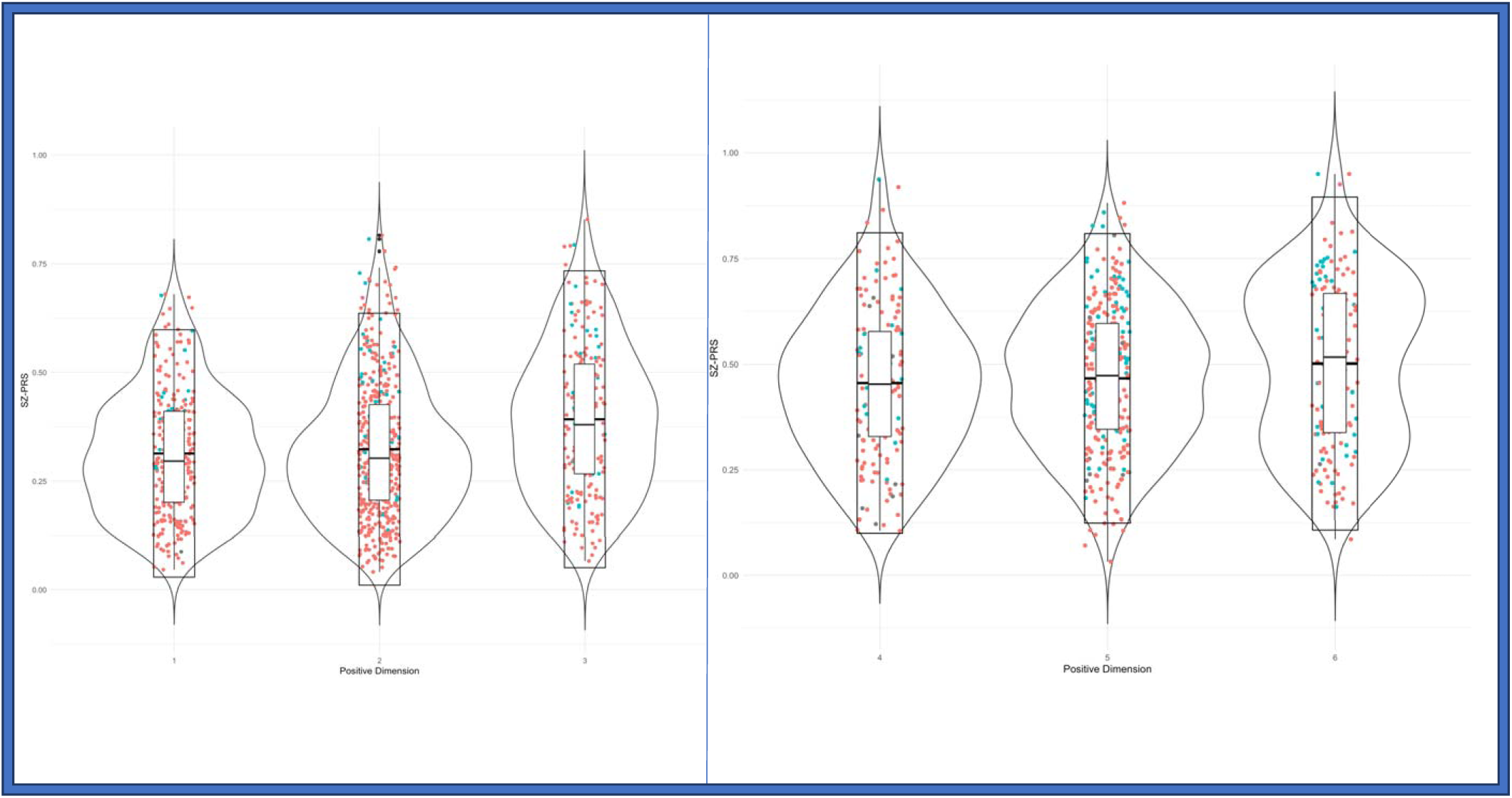
Distribution of SZ-PRS according to quantiles of psychosis in the general population and separately in FEP patients. The violin plots show the distribution of SZ-PRS in the EU-GEI sample by individuals classified according to their score at POS experience dimension and symptom dimensions, separately in population controls (left side) and FEP patients (right side) at different quantiles. In controls: (1) 0-25% psychotic experiences; (2) 25-75% psychotic experiences; and (3) 75-100% psychotic experiences. In FEP: (4) 0-25% psychotic symptoms; (5) 25-75% psychotic symptoms; and (6) 75-100% psychotic symptoms. Explanatory note: Interquartile range, 95% confidence interval, median and mean are illustrated within the bars. On each side of the bars is represented a kernel density estimation to show the distribution shape of the data. Dots indicate current cannabis use in controls and daily cannabis use in patients (red= no; green = yes)

### POS symptom dimensions by PRS and cannabis use in patients and controls

Daily cannabis use (B=0.31; 95%CI 0.11 to 0.52; p=0.002) and SZ-PRS (B=0.22; 95%CI 0.04 to 0.39; p=0.014) were independently associated with POS symptoms in patients, and this joint model improved fit over a model with SZ-PRS alone (LR chi2(1)=6.10, p = 0.01).

Similar results were found for POS psychotic experiences in controls, with main effects of current use of cannabis (B=0.26, 95%CI 0.06 to 0.46; p=0.011) and SZ-PRS (B=0.13, 95%CI 0.02 to 0.25; p=0.022) (Figure 4), and with an improvement of model fit (LR chi2(1)=6.42, p = 0.01).

**Figure 4.**
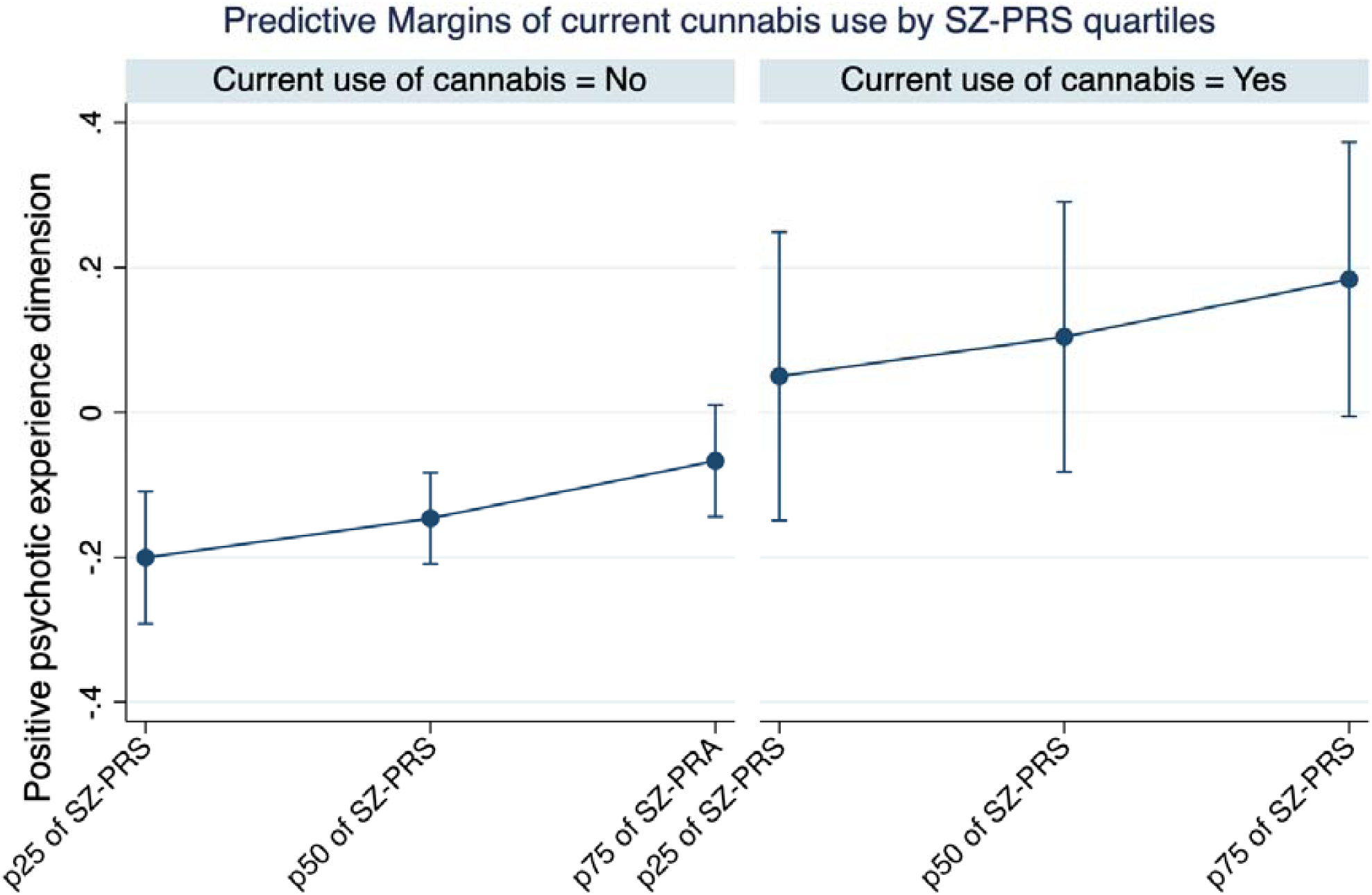
POS dimensions by SZ-PRS and cannabis use in controls. The graph presents the predicted POS symptom dimension scores at two different covariate values (cannabis use yes/no), holding SZ-PRS at 25^th^, 50^th^, and 75^th^ percentiles. Predicted values were adjusted for age, sex, and 10 ancestry PCs.

## Discussion

### Principal findings

This is the first study to investigate the combined effect of SZ-PRS and cannabis use on psychosis dimensions. We found that these two factors, independently from each other, are associated with more clinical and sub-clinical POS symptoms in both FEP patients and controls. Moreover, we found a relationship between SZ-PRS and more NEG symptoms and experiences. Finally, we did not find in our sample an association between BP-PRS and MAN symptoms; or between the combined SZ *&* BP-PRS and the G factor.

Our findings provide first evidence that in patients and controls, the latent structure of psychosis, as generated using a statistically guided approach, is valid and coheres with both SZ risk variants and cannabis use. However, any further interpretation on the applicability of these findings should take into account the small magnitude of all the detected associations.

### Comparison with previous research

Our findings extend those from previous research on the validity of psychosis symptom dimensions by ascertaining their coherence with genetic factors and cannabis use. First, under the hypothesis that psychosis symptom presentation is partly a function of SZ genetic liability, we reported an association between SZ-PRS and both POS and NEG symptom dimensions. This in line with a meta-analysis suggesting that different SZ risk loci impact on SZ clinical heterogeneity, e.g. genes related to immune system might be overrepresented for NEG, and genes related to addiction and dopamine-synapses might be overrepresented for POS (58).

Familial co-aggregation of NEG symptoms was reported in the Danish adoption study (19), in the Roscommon family study (59), in the Maudsley twin series studies (18). Genome-wide suggestive linkages with an effect on NEG symptoms have also been reported, although without reaching a significance threshold (60, 61). GWAS and PRS examinations provide good evidence of a polygenic signal for NEG (26-30). Altogether, these studies provide converging evidence that NEG has substantive heritability at least partly due to cumulative schizophrenia risk loci. The DIS dimension has also been reported as having high heritability in some studies (20), but we found no evidence of its association with SZ-PRS in our FEP sample, and we could not examine this latent construct in our controls. Speculatively, it is possible that DIS symptom differ in their lifetime v. FEP prevalence, or that genetic loci influencing DIS are different from those carrying SZ risk (20).

Second, our results on the relationship between SZ-PRS and POS are intriguing but less consistent with previous literature. Possible familial co-aggregation of POS symptoms was rarely reported (62, 63). However, a previous study observed that BP patients with higher SZ-PRS presented with more mood-incongruent POS symptoms (64), which suggests SZ-PRS has a POS modifier effect. Nevertheless, this was not confirmed by meta-analysis of PGC and GPC samples (30, 65). We may consider in interpreting our data, that the EU-GEI sample included FEP patients only, hence symptomatology rating was not confounded by antipsychotic treatment; whereas PGC and GPC samples are most chronic schizophrenia samples, where the enduring antipsychotic treatment can attenuate POS symptoms and increase NEG symptoms (i.e., secondary NEG symptoms). Moreover, various environmental factors impacting at different levels on dopaminergic activity makes it difficult to disentangle the risk variants contribution to POS symptoms over the course of SZ. From this perspective, we extend previous evidence that use of cannabis is associated with more POS symptomatology at FEP (11, 66), clarifying that this association is independent from SZ genetic risk loading.

Third, unlike our hypothesis and larger studies (29, 67), we did not report an association between BP-PRS and MAN. This may suggest that our sample is too small for BP-PRSs based on GWAS than SZ, or the true effect of BP-PRS is too small.

Fourth, we replicated in our controls the same patterns of associations as in cases between SZ-PRS and dimensions, but in the form of sub-clinical psychosis. Further, we provide novel evidence that SZ-PRS and current cannabis use are both associated with more POS psychotic experiences. It has been debated whether sub-clinical psychotic symptoms have an etiological overlap with full-blown psychosis.

Our findings support the evidence that SZ-PRS correlates with psychotic experiences (35), which in adults may be reflecting similarities with biological SZ risk factors (35). Moreover, a few SNPs reaching genome-wide significance have been recently identified for psychotic experiences, for example in *CNR2*, coding for the cannabinoid receptor type 2 (68). This suggests that further studies are needed to clarify the relationship between patterns of cannabis use and sets of genes potentially enhancing its psychotropic effects (69).

Finally, to our knowledge this is the first study examining SZ *&* BP-PRS and G factor in psychotic disorders, under the hypothesis they have a positive correlation. We report a negative finding which may be explained by G not properly reflecting general psychopathology in our FEP sample. On other hand, the G factor of psychotic experiences in controls well correlated with SZ genetic liability.

### Limitations

The following limitations suggest exercising caution when interpreting our findings.

1. We performed extensive work for defining the fine-scale population structure in a multi-ethnic sample. Certainly, having a sample of individuals from a single homogenous population would have improved the quality of the analysis, however our study has the advantage of being more representative of the real clinical practice. Most important, we included as far as possible population clusters not located in Europe but still suitable for PRS analyses, which is in line with a more general aim of not contributing to healthy disparities (70).
2. Regarding symptom ratings in patients, we used symptom dimensions from two different scales, i.e. NEG from SDS, and the other symptom dimensions from OPCRIT. In the EU-GEI study, negative symptoms were rated through the administration of SDS; moreover, exploratory factor analyses of OPCRIT in other samples showed that a hybrid DIS/NEG dimension was often obtained rather than discrete NEG and DIS dimensions (30, 71). Of note, our preliminary analysis of SZ-PRS and NEG using OPCRIT showed no nominal association (72), due, possibly, to the scarce item covariance coverage, acknowledged as a limitation in our earlier paper on symptom dimensions (10).
3. Regarding the bifactor solutions, G may be difficult to interpret and possibly overfits the data (73). Nevertheless, in our model, G improves the measurement of specific dimensions by making their score not unduly affected by the all-item covariance (10). Moreover, based on the strength of item factor loadings in our sample, G could be interpreted: 1) in patients, as combined manic-delusional symptomatology (10); 2) in controls, as a combined measure of all types of psychotic experiences (11).
4. We did not validate self-reported information on current use of cannabis with biological samples. However, this method does not allow ascertaining lifetime patterns of cannabis use (40) and is not considered a gold standard method (74). Moreover, it has been shown that self-report information on cannabis use is consistent with laboratory data (75).
5. We did not use a PRS based on GWAS of symptom dimensions, as this is currently unavailable. It is noteworthy that, genes conferring risk to a disorder (‘risk genes’) may not overlap with genes modifying symptom presentation (‘modifier genes’) (76), although it is hypothesised that there are genes with a mixed effect (58). Thus, our study answers the question whether the genetic liability for psychotic disorder explains variance of some phenotypic traits, without accounting for other possible genetic sources of that variance (i.e., the contribution of modifier genes, copy number variants, and rare SNPs).

### Implications

Most clinical and research psychiatrists still embrace Kraepelin’s nosology in the field of psychosis, despite the fact that for a century concerns have been raised related to the absence of converging validators to distinguish non-affective and affective psychotic disorders (77). We report two classes of external validators of transdiagnostic symptom dimensions, such as SZ-PRS and cannabis use. It should be born in mind that pharmacological and psychological interventions, as well as cannabis cessation and all secondary prevention strategies target particular symptoms more than the general diagnosis. Hence, our findings support the concept of a psychosis continuum.

## Data Availability

Any request has to be considered under the provision of the EU-GEI project procedures

## Acknowledgements

The work was supported by: Clinician Scientist Medical Research Council fellowship (project reference MR/M008436/1) to MDF; the National Institute for Health Research (NIHR) Collaboration for Leadership in Applied Health Research and Care South London at King’s College Hospital NHS Foundation Trust to DQ.; National Institute for Health Research (NIHR) Biomedical Research Centre for Mental Health at South London and Maudsley NHS Foundation Trust and King’s College London. The views expressed are those of the author(s) and not necessarily those of the NHS, the NIHR or the Department of Health and Social Care.

The EU-GEI Project is funded by the European Community’s Seventh Framework Programme under grant agreement No. HEALTH-F2-2010-241909 (Project EU-GEI). The Brazilian study was funded by the São Paulo Research Foundation under grant number 2012/0417-0.

Funders were not involved in design and conduct of the study; collection, management, analysis and interpretation of the data; preparation, review or approval of the manuscript, and decision to submit the manuscript for publication.

## Appendix

^*‡*^**EU-GEI group authorship includes:**

Kathryn Hubbard^1^, Stephanie Beards^1^, Simona A. Stilo^2^, Mara Parellada^3^, David Fraguas^3^, Marta Rapado Castro^3^, Álvaro Andreu-Bernabeu^3^, Gonzalo López^3^, Mario Matteis^3^, Emiliano González^3^, Manuel Durán-Cutilla^3^, Covadonga M. Díaz-Caneja^3^, Pedro Cuadrado^4^, José Juan Rodríguez Solano^5^, Angel Carracedo^6^, Javier Costas^6^, Emilio Sánchez^7^, Bibiana Cabrera^8^, Esther Lorente-Rovira^9^, Paz Garcia-Portilla^10^, Estela Jiménez-López^11^, Nathalie Franke^12^, Daniella van Dam^12^, Fabian Termorshuizen^13,14^, Nathalie Franke^13^, Elsje van der Ven^13,14^, Elles Messchaart^14^, Marion Leboyer^15,16,17,18^, Franck Schu rhoff^15,16,17,18^, Stéphane Jamain^16,17,18^, Grégoire Baudin^15,16^, Aziz Ferchiou^15,16^, Baptiste Pignon^15,16,18^, Jean-Romain Richard^16,18^, Thomas Charpeaud^18,19,21^, Anne-Marie Tronche^18,19,21^, Flora Frijda^22^, Giovanna Marrazzo^23^, Lucia Sideli^22^, Crocettarachele Sartorio^22,23^, Fabio Seminerio^22^, Camila Marcelino Loureiro^24,25^, Rosana Shuhama^24,25^, Mirella Ruggeri^26^, Chiara Bonetto^26^, Doriana Cristofalo^26^, Domenico Berardi^27^, Marco Seri^27^, Giuseppe D’Andrea^27^.

**Affiliations**

^1^ Department of Health Service and Population Research, Institute of Psychiatry, King’s College London, De Crespigny Park, Denmark Hill, London, United Kingdom, SE5 8AF

^2^ Department of Psychosis Studies, Institute of Psychiatry, King’s College London, De Crespigny Park, Denmark Hill, London, United Kingdom SE5 8AF

^3^ Department of Child and Adolescent Psychiatry, Hospital General Universitario Gregorio Marañón, School of Medicine, Universidad Complutense, IiSGM (CIBERSAM), C/Doctor Esquerdo 46, 28007 Madrid, Spain

^4^ Villa de Vallecas Mental Health Department, Villa de Vallecas Mental Health Centre, Hospital Universitario Infanta Leonor / Hospital Virgen de la Torre, C/San Claudio 154, 28038 Madrid, Spain

^5^ Puente de Vallecas Mental Health Department, Hospital Universitario Infanta Leonor / Hospital Virgen de la Torre, Centro de Salud Mental Puente de Vallecas, C/Peña Gorbea 4, 28018 Madrid, Spain

^6^ Fundación Pública Galega de Medicina Xenómica, Hospital Clínico Universitario, Choupana s/n, 15782 Santiago de Compostela, Spain

^7^ Department of Psychiatry, Hospital General Universitario Gregorio Marañón, School of Medicine, Universidad Complutense, IiSGM (CIBERSAM), C/Doctor Esquerdo 46, 28007 Madrid, Spain

^8^ Department of Psychiatry, Hospital Clinic, IDIBAPS, Centro de Investigación Biomédica en Red de Salud Mental (CIBERSAM), Universidad de Barcelona, C/Villarroel 170, escalera 9, planta 6, 08036 Barcelona, Spain

^9^ Department of Psychiatry, School of Medicine, Universidad de Valencia, Centro de Investigación Biomédica en Red de Salud Mental (CIBERSAM), C/Avda. Blasco Ibáñez 15, 46010 Valencia, Spain

^10^ Department of Medicine, Psychiatry Area, School of Medicine, Universidad de Oviedo, Centro de Investigación Biomédica en Red de Salud Mental (CIBERSAM), C/Julián Clavería s/n, 33006 Oviedo, Spain

^11^ Department of Psychiatry, Servicio de Psiquiatría Hospital “Virgen de la Luz”, C/Hermandad de Donantes de Sangre, 16002 Cuenca, Spain

^12^ Department of Psychiatry, Early Psychosis Section, Academic Medical Centre, University of Amsterdam, Meibergdreef 5, 1105 AZ Amsterdam, The Netherlands

^13^ Rivierduinen Centre for Mental Health, Leiden, Sandifortdreef 19, 2333 ZZ Leiden, The Netherlands

^14^ Department of Psychiatry and Neuropsychology, School for Mental Health and Neuroscience, South Limburg Mental Health Research and Teaching Network, Maastricht University Medical Centre, P.O. Box 616, 6200 MD Maastricht, The Netherlands

^15^ AP-HP, Groupe Hospitalier “Mondor”, Pôle de Psychiatrie, 51 Avenue de Maréchal de Lattre de Tassigny, 94010 Créteil, France

^16^ INSERM, U955, Equipe 15, 51 Avenue de Maréchal de Lattre de Tassigny, 94010 Créteil, France

^17^ Faculté de Médecine, Université Paris-Est, 51 Avenue de Maréchal de Lattre de Tassigny, 94010 Créteil, France

^18^ Fondation Fondamental, 40 Rue de Mesly, 94000 Créteil, France

^19^ CMP B CHU, BP 69, 63003 Clermont Ferrand, Cedex 1, France

^20^ Etablissement Public de Santé Maison Blanche, Paris, France

^21^ Université Clermont Auvergne, EA 7280, Clermont-Ferrand, 63000, France

^22^ Department of Experimental Biomedicine and Clinical Neuroscience, Section of Psychiatry, University of Palermo, Via G. La Loggia n.1, 90129 Palermo, Italy

^23^ Unit of Psychiatry, “P. Giaccone” General Hospital, Via G. La Loggia n.1, 90129 Palermo, Italy

^24^ Departamento de Neurociências e Ciencias do Comportamento, Faculdade de Medicina de Ribeirão Preto, Universidade de São Paulo, Av. Bandeirantes, 3900 -Monte Alegre-CEP 14049-900, Ribeirão Preto, SP, Brasil

^25^ Núcleo de Pesquina em Saúde Mental Populacional, Universidade de São Paulo, Avenida Doutor Arnaldo 455, CEP 01246-903, SP, Brasil

^26^ Section of Psychiatry, Department of Neuroscience, Biomedicine and Movement, University of Verona, Piazzale L.A. Scuro 10, 37134 Verona, Italy

^27^ Department of Medical and Surgical Science, Psychiatry Unit, Alma Mater Studiorum Università di Bologna, Viale Pepoli 5, 40126 Bologna, Italy

